# Daily Rhythms in Blood Glucose: Time-of-Day Forecasts in Type 2 Diabetes

**DOI:** 10.1101/2025.06.02.25328803

**Authors:** Nicolai Peder Bülow Pedersen, Tanja Kortsen Bugajski, J. Eduardo Vera-Valdés, Stine Hangaard Casper, Morten Hasselstrøm Jensen, Peter Vestergaard, Thomas Kronborg

## Abstract

**Background:** Accurate and interpretable forecasting of blood glucose levels is critical for effective management of Type 2 diabetes. While complex machine learning models offer high predictive accuracy, their opacity often limits clinical applicability. This study investigates the performance of a simple, interpretable reference model: the time-of-day mean forecast.

**Method:** The proposed approach divides each 24-hour period into discrete time sequences and, for each sequence, computes the mean glucose value across previous days. This methodology captures intra-day regularities in glucose dynamics and implicitly accounts for circadian influences, such as variations in insulin sensitivity and hepatic glucose production.

**Results:** The model reflects intra-day glucose patterns and identifies clinically relevant periods of elevated variability, such as the postprandial and nocturnal windows. Forecasting performance improves with increased temporal granularity: in 91.84% of the individuals, at least one finer bin size outperformed the naïve baseline. Where, 51% achieved optimal performance using the highest resolution with a 5-minute bin size. Compared to the naïve approach, the 5-minute bin size reduced mean squared error by an average of 12.2%.

**Conclusions:** We have justified the time-of-day approach using a simple mean forecast model, showing that aligning prediction windows with time-of-day patterns enhances forecast accuracy. Building on this foundation, the time-of-day mean forecast serves as a practical benchmark. Future work should explore more complex models that incorporate individual covariates and dynamic temporal dependencies, while maintaining interpretability using the described temporal structure.

## Introduction

Numerous studies have demonstrated that glucose regulation follows a circadian rhythm, in Type 2 Diabetes (T2D), glucose levels exhibit daily fluctuations, including well-documented postprandial excursions and variations (1–4). These recurring patterns suggest that, even without complex physiological modelling, it is possible to estimate typical glucose behaviour at specific times of day. For instance, predictable postprandial spikes often occur after breakfast, lunch, and dinner, while lower levels are typically observed overnight or between meals. Capturing this regularity may serve as a strong baseline for glucose forecasting (5,6).

In this study, we investigate the performance of a simple yet interpretable forecasting model: the time-of-day mean forecast. The approach partitions the 24-hour day into discrete time sequences and, for each sequence, calculates the average glucose level from previous days. Under the assumption that the intra-day pattern of glucose is stable across days, this average provides an estimate of the expected glucose level at that time.

Unlike more complex models requiring substantial data and computational resources, the time-of-day mean forecast is straightforward to implement and easy to explain. Its transparency makes it particularly suitable for clinical settings, where interpretability is crucial for decision-making.

By aligning predictions with specific times of day, the method naturally incorporates circadian influences on glucose metabolism, including variations in insulin sensitivity and hepatic glucose production. This allows us to highlight periods of higher variability or glycaemic risk, such as the postprandial window or late evening hours.

Time-of-day averaging methods are widely used in domains such as finance, where intraday trading patterns and volatility are often analysed using historical averages at fixed time intervals. These methods help capture systematic daily trends while providing interpretable insights into uncertainty and risk (7). By adapting a similar methodology, we leverage its proven strengths in capturing regular temporal patterns and estimating time-specific variability, now applied to the clinical context of Continuous Glucose Monitoring (CGM) data.

This approach, paired with the quantification of prediction uncertainty via sample variance, offers a simple yet powerful method for capturing intra-day glucose variability at specific times. Not only does it quantify the uncertainty in predictions, but it also provides valuable clinical insights, laying a rigorous foundation for evaluating the performance of more sophisticated predictive models.

## Method

### Data Source

The data used in this study originated from the trial “Diabetes teleMonitoring of patients in insulin Therapy (DiaMonT)” as part of the “Adherence through Cloud-based Personalized Treatment for Type 2 Diabetes (ADAPT-T2D) consortium” (8). The study was a randomized controlled trial that enrolled 329 participants with insulin-treated type 2 diabetes for a study period of 90 days. The participants in the intervention arm were equipped with a Dexcom G6 CGM sensor connected to their smartphone throughout the entire study period, whereas the control arm wore a blinded sensor for the first and last 20 days of the study. The CGM sensor provided estimates of the blood glucose values in 5-minute intervals throughout the day and thus great insights into the glucose fluctuations to address the aim of this present study.

### Missing Values

Missing values and measurement inconsistencies, such as duplicate glucose readings (likely due to synchronisation errors or logging redundancies), were identified and marked as *NaN*. This explicit marking ensures that any gaps and errors are appropriately handled during data preprocessing.

### Alignment

We included 147 out of 329 participants who met the criterion of at least 90% data completeness over the 90-day trial period. All time series were aligned to begin at 00:00:00 on the first full day following participant inclusion. Any measurements recorded prior to this time were excluded to ensure consistency across participants and enable valid time-of-day comparisons. To maintain uniformity in analysis, all time series were truncated to 83 days; the maximum common duration across participants.

### Partitioning Approach

The novelty of our design is that the time series is reorganised into smaller, more manageable sequences. Specifically, each time series is partitioned into 288 sequences, each corresponding to a unique 5-minute timestamp within a 24-hour period (e.g., 00:00:00, 00:05:00, 00:10:00, etc.). For example, the first sequence concatenates all glucose values recorded at 00:00:00 across all days, the second includes values from 00:05:00, and so on. This structure is illustrated in Table 1, where each column represents a distinct sequence identified by its timestamp.

**Table 1:** Time-of-day partitioning

| Day | Sequence 1 | Sequence 2 | Sequence 3 | ... | Sequence 288 |
| --- | --- | --- | --- | --- | --- |
| Day 1 | 00:00:00 | 00:05:00 | 00:10:00 | ... | 23:55:00 |
| Day 2 | 00:00:00 | 00:05:00 | 00:10:00 | ... | 23:55:00 |
| Day 3 | 00:00:00 | 00:05:00 | 00:10:00 | ... | 23:55:00 |
| ... | ... | ... | ... | ... | ... |
| Day 83 | 00:00:00 | 00:05:00 | 00:10:00 | ... | 23:55:00 |

This partitioning isolates time-specific variation in Blood Glucose Levels (BGLs) and facilitates the analysis of circadian patterns in glucose regulation. The core idea is that increases due to physiological factors (e.g., meals, exercise) can be captured more effectively within these time-specific sequences. People tend to have regular daily routines, such as eating and sleeping, which can be reflected in their glucose levels at specific times of day. This restructuring facilitates alignment of daily rhythms across multiple days, providing a consistent framework for time-of-day analysis. Missing values within each sequence are estimated and filled using linear interpolation.

## Method

### Methodology

By partitioning the data into specific time intervals, we can capture these predictable patterns and compute an estimate of the expected glucose level at each time interval (6,9).

The theoretical rationale behind the method is that the glucose value *G* at some time *t*, on any given day, can be thought of as a sample from the same distribution as the glucose values at (the same) time *t*, on all previous days. The assumption that glucose patterns at each time are stationary allows us to estimate this underlying distribution using the average glucose level at that time across multiple days.

The method leverages time-specific regularities and behavioural routines embedded in CGM data, offering a transparent and data-driven benchmark for evaluating more complex forecasting models in the future. As a benchmark, we consider a naïve implementation of the mean forecast without partitioning. Then, we will explore the impact of partitioning the data into time-specific sequences by increasing the bin size (i.e., the aggregated bin size used for prediction) to study potential improvements in predictive performance.

### Statistical Interpretation of the Mean Forecast

Let *G*_*n,t*_ denote the glucose value at time *t* on day *n*, for *n* = 1, …, *N*, where *N* is the total number of days used in the forecast. The time-of-day mean forecast is defined as the sample average:

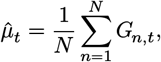

which consistently estimates the expected glucose level *μ*_*t*_ = 𝔼[*G*_*n,t*_] at time *t* under temporal stationarity.

### Stationarity and Autocorrelation

We assume that the distribution of glucose at each fixed time of day *t* is stationary across days with a constant mean *μ*_*t*_, but with autocorrelated and potentially heteroskedastic residuals. Specifically, we model:

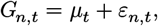

where *ε*_*n,t*_ is a zero-mean error term that exhibits serial correlation in time and may exhibit varying in scale. That is, the errors at different times of day may not be independent and may have different variances.

Under these assumptions, the sample mean 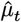 remains a consistent estimator of *μ*_*t*_, and the effective variance of 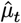 is estimated via a heteroskedasticity and autocorrelation consistent (HAC) estimator:

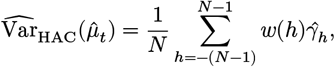

where *w*(*h*) is a kernel weight and 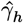 is the empirical autocovariance at lag *h*:

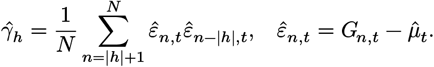

The HAC estimator provides consistent variance estimates even when errors exhibit heteroskedasticity and dependence (10).

### Large Sample Properties

Under general heteroskedasticity and weak temporal dependence across days, we can appeal to asymptotic results to justify inference on the time-of-day mean estimate 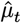

The weak Law of Large Numbers (LLN) ensures:

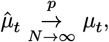

and the Central Limit Theorem (CLT) under weak dependence gives:

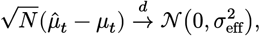

where 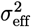 is the long-run variance incorporating both heteroskedasticity and autocorrelation, estimated using a HAC estimator.

In summary, the LLN ensures that 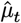 consistently estimates the true mean *μ*_*t*_ as the number of days *N* grows large. Meanwhile, the CLT justifies approximating the distribution of 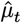 by a normal distribution with variance 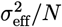, accounting for heteroskedasticity and autocorrelation. This supports valid inference and uncertainty quantification using HAC estimators.

### Forecasting Uncertainty

We define a time-of-day prediction interval (PI) for *Ĝ*_*t*_ using the estimated mean and variance:

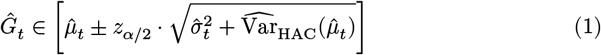

where 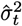 is the sample variance at time *t*:

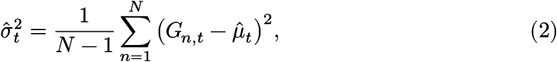

and *z*_*α*/2_ is the (1 − *α*/2)-quantile of the standard normal distribution. For example, setting *α* = 0.05 corresponds to a 95% prediction interval with *z*_0.025_ ≈ 1.96. The total uncertainty combines across-day variability and estimation error under general heteroskedasticity.

### Interpretation

- **Wider prediction intervals** reflect elevated volatility or uncertainty due to physiological or behavioural fluctuations, such as meal timing or physical activity.
- **Narrower intervals** suggest more stable, predictable glucose dynamics at that time of day.

Together, 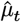 and the corresponding interval provide a robust and interpretable forecast for glucose levels, accommodating realistic assumptions of heteroskedastic and autocorrelated errors across days.

## Results

To illustrate the range of inter-individual variability in glucose dynamics, we present the CGM profiles of the subjects with the highest (HV) and lowest (LV) variance in their time series. In Figure 1, the full CGM time series showcases the broad temporal patterns for each subject, while the **Sequence 288** view highlights glucose fluctuations at a specific time of day across multiple days (as defined in Table 1).

**Figure 1.**
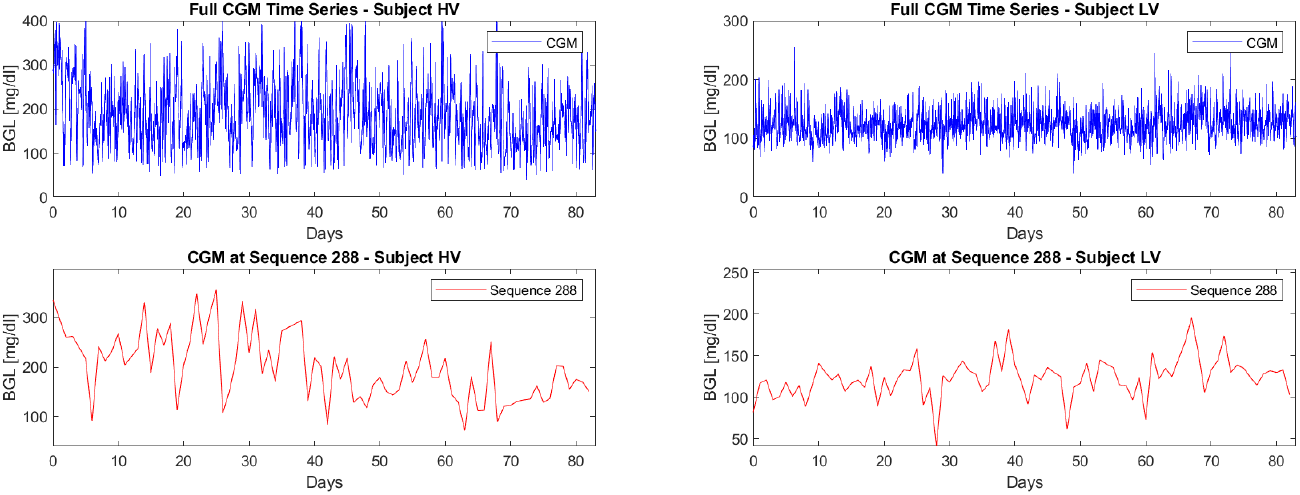
Glucose profiles of subjects with highest and lowest variance.

### Naïve Implementation of the Mean Forecast

We begin with a naïve implementation of the mean forecast over a seven-day period. The plot in Figure 2 illustrates BGLs over time, with the PI, defined by Equation 1, indicating the uncertainty surrounding the forecast.

**Figure 2.**
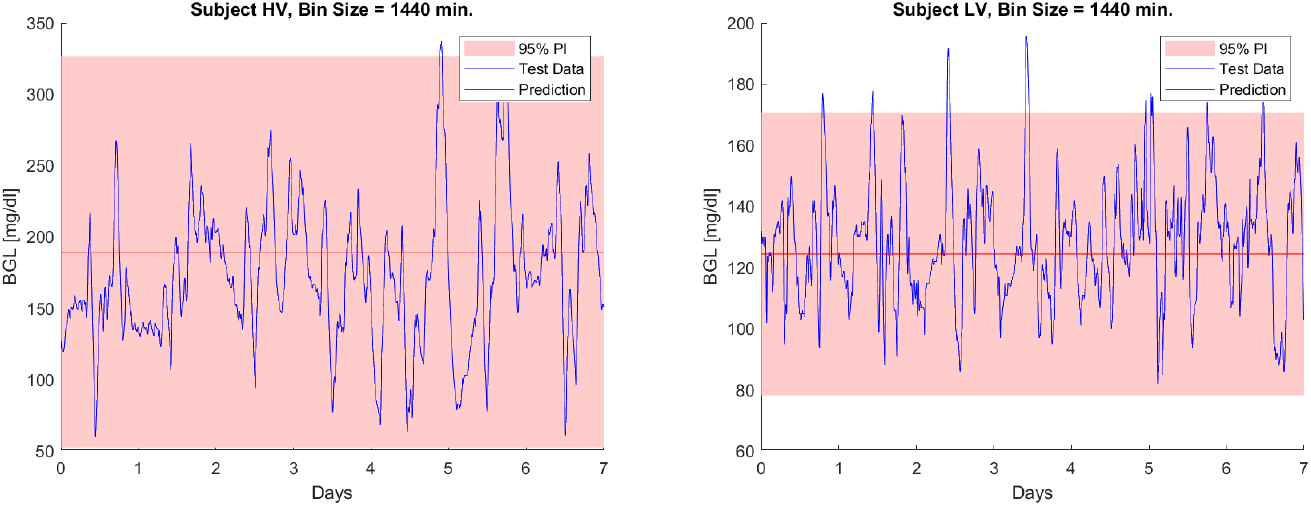
Naïve Implementation of the Mean Forecast of subjects with highest and lowest variance.

The plot demonstrates wide PIs, indicating high uncertainty and poor explanatory power. Such broad intervals hinder the identification of periods of elevated or reduced BGLs, making them unreliable for personalised treatment. This reflects the limitations of the naïve implementation, which does not account for temporal dependencies or individual variability.

### Increasing the Bin Size

We increase the bin size to 6 hours to assess potential improvements in prediction accuracy, as illustrated in Figure 3.

**Figure 3.**
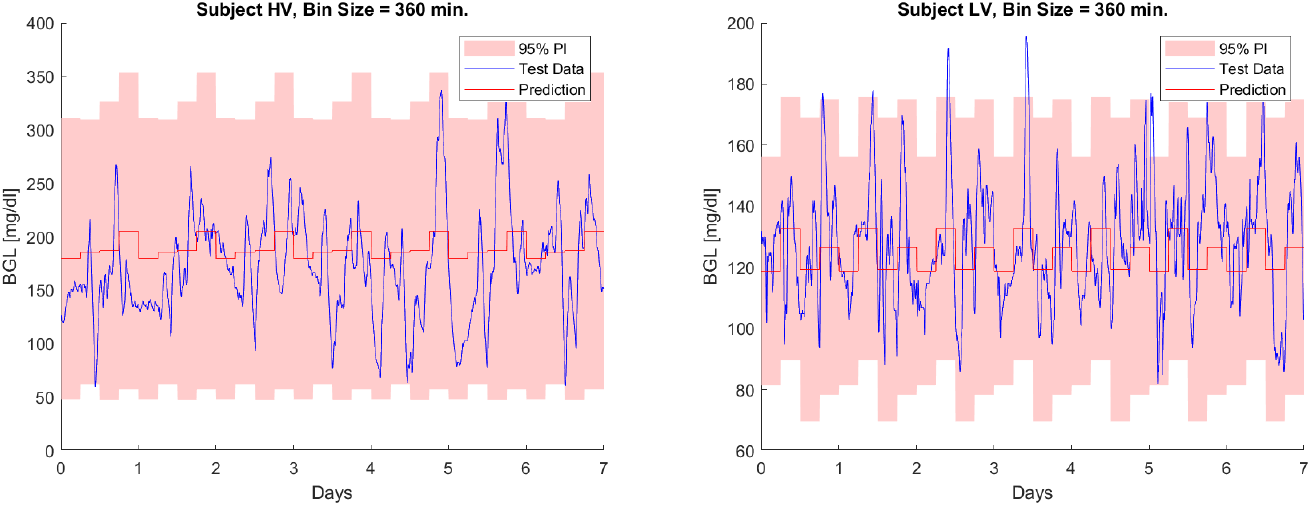
Glucose levels over time with 6 hour bin size of subjects with highest and lowest variance.

The plot demonstrates that the 6-hour mean forecast captures periods of rising or falling glucose levels throughout the day, rather than predicting a fixed value for the entire day. This dynamic prediction can facilitate the detection of anomalous glucose measurements that deviate significantly from a participant’s usual values, thereby enabling more personalised and timely treatment.

We further increase the temporal resolution by progressively reducing the bin size to one hour, 30 minutes, 15 minutes, and finally to five minutes, which corresponds to the original sampling rate. This progression is illustrated for subject (HV) in Figure 4.

**Figure 4.**
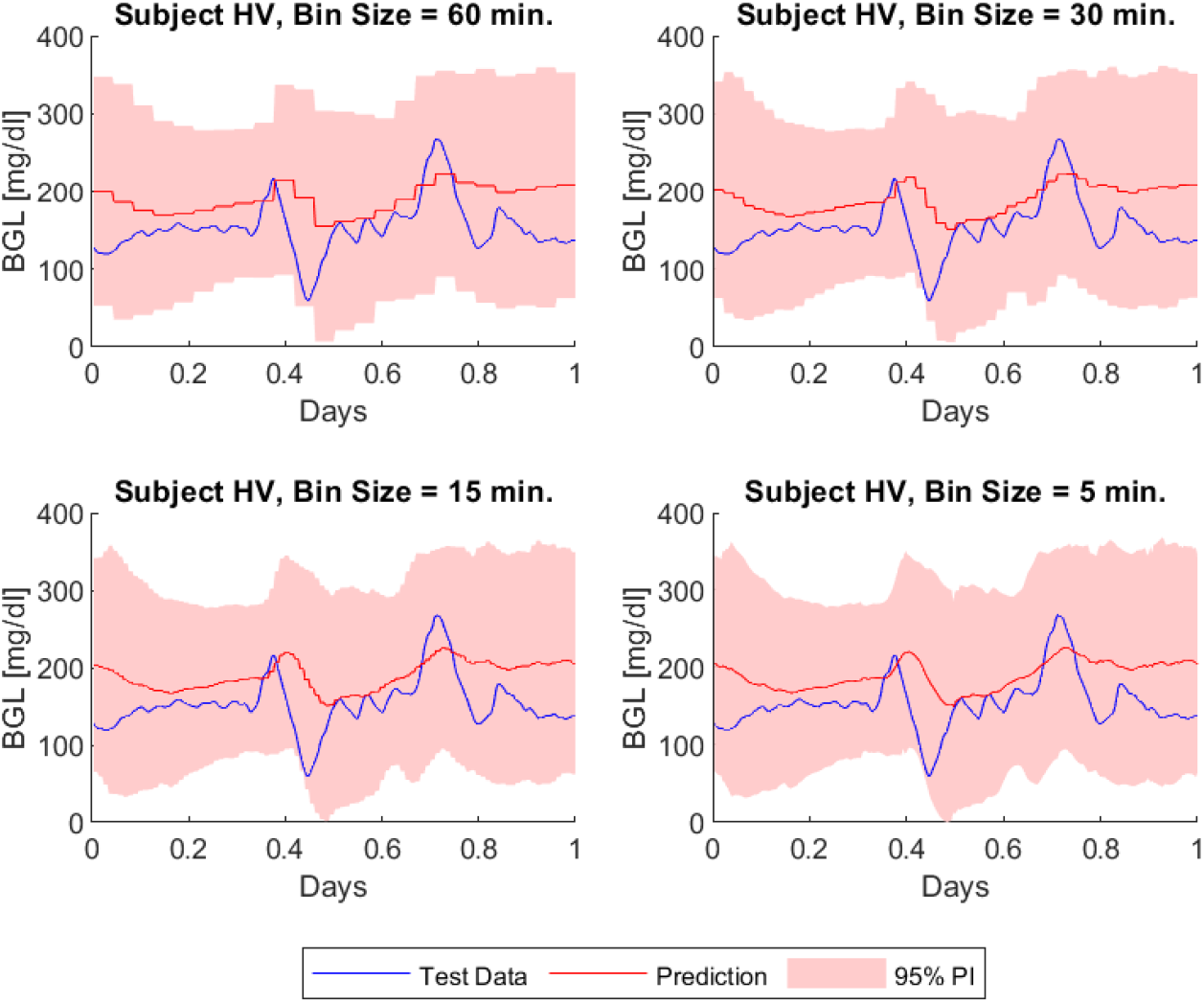
Glucose levels over time with varying bin size (Subject with high variance). And for subject (LV) in Figure 5.

**Figure 5.**
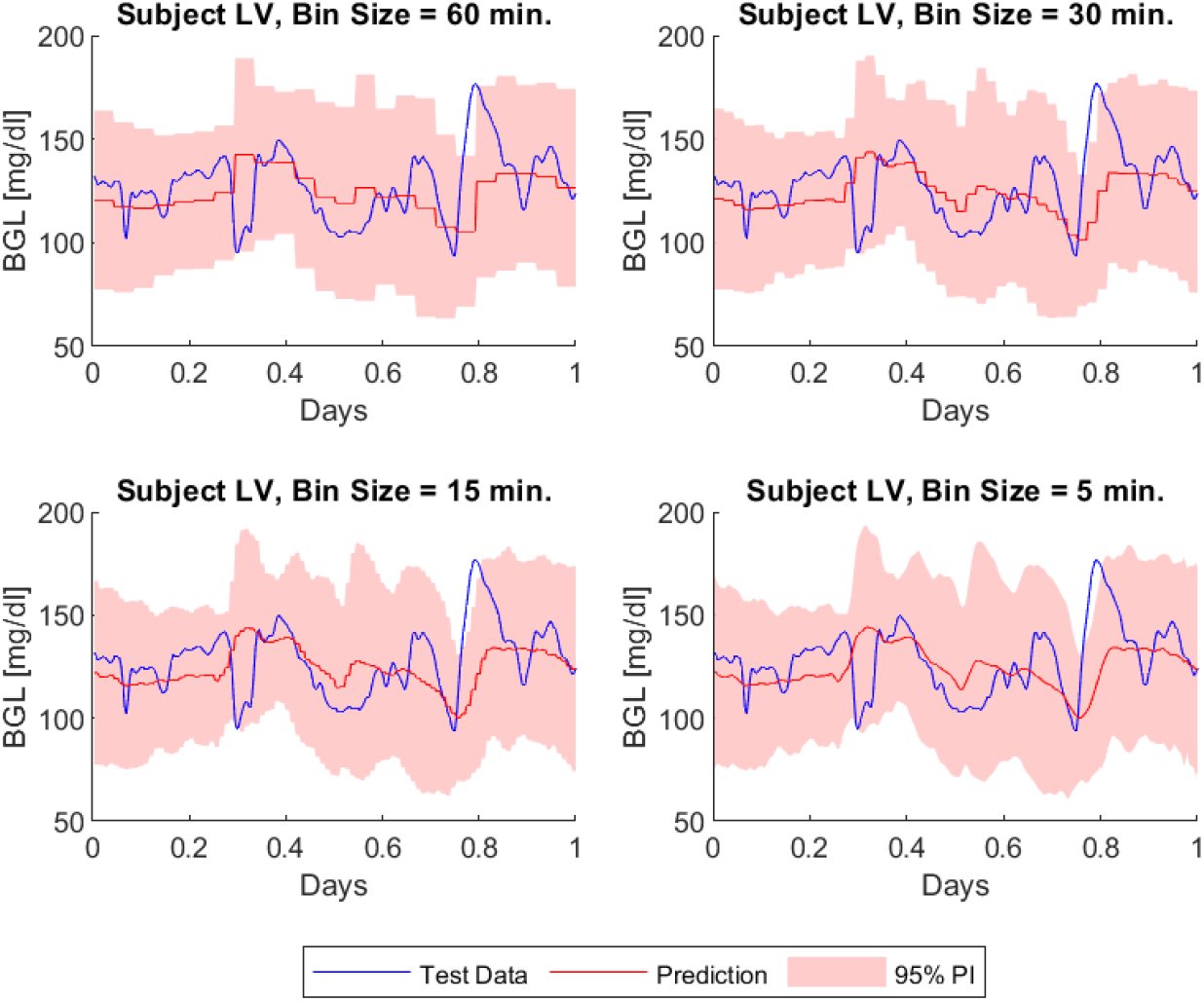
Glucose levels over time with varying bin size (Subject with low variance).

Detailed results across different bin sizes, are presented in Table 2

**Table 2:** MSE of BGLs by bin size for subject (HV) and Subject (LV), and the average across all subjects as baseline. In parentheses, the percentage change in MSE relative to the naïve implementation is shown.

| Bin Size | Subject (HV) |  | Subject (LV) |  |
| --- | --- | --- | --- | --- |
| | MSE | ΔMSE (%) | MSE | ΔMSE (%) |
| 5 min | **2094.6** | -21.7 | 353.96 | -9.2 |
| 15 min | 2095.5 | -21.6 | 354.09 | -9.2 |
| 30 min | 2098.7 | -21.5 | 354.49 | -9.1 |
| 60 min | 2112.7 | -21.0 | 357.30 | -8.4 |
| 3 hours | 2275.0 | -14.9 | 359.56 | -7.8 |
| 4 hours | 2258.0 | -15.5 | 365.98 | -6.2 |
| 6 hours | 2455.7 | -8.2 | **331.16** | -15.1 |
| 8 hours | 2256.3 | -15.6 | 389.27 | -0.2 |
| 12 hours | 2505.9 | -6.3 | 391.28 | 0.3 |
| 24 hours | 2673.7 | ref | 390.00 | ref |

The table presents the mean squared error (MSE) of glucose forecasts across various bin sizes for two representative subjects: Subject (HV), who exhibited the highest variance in their CGM time series, and Subject (LV), who exhibited the lowest variance. These two subjects were selected to illustrate the extremes of inter-individual variability in glucose dynamics. By evaluating how MSE changes with bin size, the table highlights the impact of temporal aggregation on prediction accuracy for subjects with differing levels of glycaemic variability. The corresponding percentage change in MSE relative to the baseline 24-hour bin size (naïve implementation) provides a comparative view of forecasting improvement or decline.

In Table 3, the average MSE across all subjects and the corresponding sample standard deviation (SD) given by Equation 2 are shown to quantify overall forecasting performance and inter-subject variability at each bin size.

**Table 3:** Average MSE and Sample SD.

| Bin Size | Average MSE | ΔMSE (%) | Sample SD |
| --- | --- | --- | --- |
| 5 min | **1793.3** | −12.2 | 1313.1 |
| 15 min | 1793.5 | −12.2 | 1312.9 |
| 30 min | 1794.8 | −12.1 | 1312.5 |
| 60 min | 1799.7 | −11.9 | 1311.6 |
| 3 hours | 1834.9 | −10.2 | 1308.3 |
| 4 hours | 1851.5 | −9.4 | **1306.5** |
| 6 hours | 1890.9 | −7.4 | 1307.3 |
| 8 hours | 1908.0 | −6.6 | 1306.9 |
| 12 hours | 1958.7 | −4.1 | 1332.3 |
| 24 hours | 2042.9 | ref | 1361.1 |

The results demonstrate that MSE generally decreases as the bin size decreases, indicating improved forecast accuracy at finer temporal resolutions. Subject (HV) consistently exhibits higher MSE values compared to Subject (LV) and the overall average, reflecting greater individual variability and forecast error. The sample SD of the prediction errors remains relatively stable across bin sizes, with a slight reduction at mid-range intervals (3–8 hours), suggesting more consistent predictions; however, SD increases again at larger bin sizes, indicating reduced precision due to over-aggregation. Even though the MSE is reduced when decreasing the bin size, the individual prediction errors vary considerably, indicating that, while the model may be accurate on average, it sometimes makes large errors, reducing reliability in specific instances. This variability highlights the importance of approaches that can account for individual differences.

To evaluate overall performance, Figure 6 shows the percentage change in MSE relative to the naïve forecast, plotted against each participant’s glucose variance. For each participant, each bin size is represented by a different colour, with the naïve implementation in red as a baseline plotted at zero. Markers above the baseline represent cases where MSE decreased as the bin size increased, while those below the baseline indicate improvements with smaller bin sizes. This illustrates how forecast accuracy relates to both model resolution and individual variability. The trend reveals that smaller bin sizes, especially the 5-minute resolution, consistently yield lower MSE, indicating more accurate forecasts. This supports the idea that highresolution models better capture short-term physiological fluctuations. The accompanying pie chart underscores this finding: for 51% of the subjects, the 5-minute bin size resulted in the lowest MSE across all resolutions, highlighting the benefit of fine-grained temporal modeling in glucose prediction.

**Figure 6.**
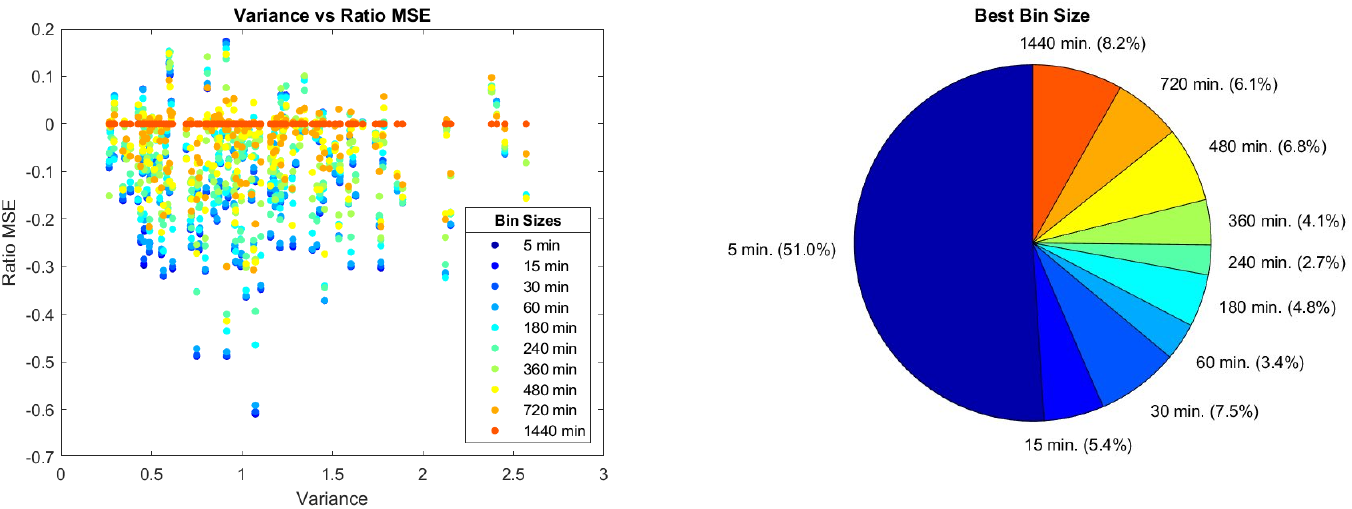
Variance vs. Ratio MSE and pie chart

Overall, we find that for 135 out of 147 subjects (91.84%), there exists a bin size that improves forecast accuracy compared to the naïve implementation. In particular, 51% of subjects achieve their lowest MSE at the 5-minute bin size, which corresponds to the smallest temporal resolution and the sampling rate. The MSE is on average lower for the 5-minute bin size, with an average reduction of 12.2% against the naïve implementation. This improvement is particularly pronounced for subjects with higher variance, where the MSE reduction reaches 21.7% for Subject (HV). This variability-sensitive improvement suggests that forecast accuracy can be enhanced by adjusting temporal resolution to individual variability levels.

## Discussion

The mean forecast provides the simplest possible implementation of our time-of-day modelling framework, offering a direct and interpretable way to summarise glucose dynamics by averaging values across days at each time point. This basic structure effectively captures daily rhythmicity without requiring complex assumptions or parameter tuning. As such, it serves as a natural and informative benchmark. However, important factors influencing glucose dynamics; such as meal timing, physical activity, medication, and stress, were not incorporated into the model. This omission limits the interpretability of forecasts and reduces the potential for personalisation tailored to individual lifestyle or physiological changes.

While the average MSE decreases with smaller bin sizes, the Sample SD remains consistently high, exceeding 1300 across all bin sizes. This indicates considerable variability in individual prediction errors. Despite improved average performance, the model may still generate large errors in some cases, which could compromise its reliability in clinical decision-making, thus, highlighting the limitations of a uniform treatment approach, supporting the need for personalised strategies. Tailoring predictions to individual patient profiles may help reduce uncertainty and improve glycaemic control, thereby enhancing clinical outcomes.

The time-of-day mean forecast is a simple approach that assumes stationarity and does not account for dynamic temporal dependencies or individual covariates, which restricts its predictive accuracy. Furthermore, this statistical model is descriptive rather than mechanistic, so it cannot explain the causes of glucose fluctuations or predict responses to interventions.

Our primary purpose was to demonstrate how forecast accuracy improves as the bin size decreases, particularly when the forecast resolution matches the data sampling rate. Our results show that reducing the forecast bin size generally improves accuracy, highlighting the importance of temporal resolution in capturing meaningful physiological variation. Importantly, we found that 91.84% of subjects benefit from this high-resolution approach, with 51% achieving their lowest MSE at the 5-minute resolution. Moreover, individuals with greater glucose variability experience the most pronounced improvements, suggesting that even a simple, high-frequency forecast can provide meaningful personalisation. This reinforces the broader point that aligning model granularity with individual dynamics is critical. Despite its simplicity, the mean forecast offers a valuable benchmark for evaluating more sophisticated prediction methods.

## Conclusion

The mean forecast offers a simple yet effective baseline for predicting glucose levels, capturing daily rhythms by averaging historical data at each time point. While it lacks contextual or dynamic features, our results show that its performance improves markedly at finer temporal resolutions, particularly when aligned with the data’s sampling rate.

As a transparent and interpretable model, it provides a valuable point of comparison for evaluating more sophisticated forecasting methods. Future models should build on this foundation by incorporating contextual information, such as meals, activity, medication, and stress, and by modelling individual dynamics over time. These advances could enable more accurate and personalised glucose predictions.

Despite its limitations, the mean forecast remains a robust starting point for glucose modelling and a meaningful benchmark for future improvements.

## Data Availability

The data utilized (DiaMont study) in this study are not publicly available due to the inclusion of sensitive patient information, which is subject to strict confidentiality and privacy regulations. Access to the data is restricted to ensure compliance with ethical guidelines and to protect patient privacy. Requests for additional information or collaboration may be considered on a case-by-case basis, subject to appropriate ethical approval and data-sharing agreements.

## Contact Information

**Nicolai Peder Bülow Pedersen**

Department of Mathematical Sciences

Aalborg University

Thomas Manns Vej 23, 9220 Aalborg

**Tanja Kortsen Bugajski**

Department of Mathematical Sciences

Aalborg University

Thomas Manns Vej 23, 9220 Aalborg

**J. Eduardo Vera-Valdés, PhD**

Department of Mathematical Sciences

Aalborg University

Thomas Manns Vej 23, 9220 Aalborg

**Stine Hangaard Casper, PhD**

Department of Health Science and Technology

Aalborg University

Gistrup, 9260 Aalborg, Denmark

**Morten Hasselstrøm Jensen, PhD**

Department of Health Science and Technology

Aalborg University

Gistrup, 9260 Aalborg, Denmark

**Peter Vestergaard, PhD**

Steno Diabetes Center North Denmark

Aalborg University Hospital

Gistrup, 9260 Aalborg, Denmark

**Thomas Kronborg, PhD**

Department of Health Science and Technology

Aalborg University

Gistrup, 9260 Aalborg, Denmark

### Abbreviations

T2D: Type 2 Diabetes
CGM: Continuous Glucose Monitoring
DiaMonT: Diabetes teleMonitoring of patients in insulin Therapy
ADAPT-T2D: Adherence through Cloud-based Personalized Treatment for Type 2 Diabetes
BGL: Blood Glucose Level
HAC: Heteroskedasticity and Autocorrelation Consistent
LLN: Law of Large Numbers
CLT: Central Limit Theorem
PI: Prediction Interval
HV: High Vaiance
LV: Low Variance
MSE: Mean Squared Error
SD: Standard Deviation

## Figures and Tables

6 figures, 3 tables.

## Acknowledgements

We would like to thank the ADAPT-T2D (Adherence through cloud-based Personalised Treatment for Type 2-Diabetes) consortium for the opportunity to conduct this project and for providing access to the data.

## CRediT authorship contribution statement

**Nicolai Peder Bülow Pedersen**: Conceptualization, Methodology, Software, Formal analysis, Writing – original draft. **Tanja Kortsen Bugajski**: Conceptualization, Methodology, Software, Formal analysis, Writing – original draft. **J. Eduardo Vera-Valdés**: Conceptualization, Methodology, Formal analysis, Supervision, Writing – original draft. **Stine Hangaard Casper**: Data Curation, Writing – review & editing. **Morten Hasselstrøm Jensen**: Data Curation, Writing – review & editing. **Peter Vestergaard**: Data Curation, Writing – review & editing. **Thomas Kronborg**: Data Curation, Writing – review & editing.

